# Using Mobile Phone Infrastructure Provides Evidence of a Reduction in Diagnostic Turnaround Times and Improved HIV Viral Load Monitoring in Malawi

**DOI:** 10.1101/2025.01.22.25320358

**Authors:** Rachel Haggard, Christopher Mwase, Brandon Klyn, Lynn Metz, Tyler Smith, Hannah Cooper, Brown Chiwandira, Linley Chewere, Dylan Green

## Abstract

**Background:** Malawi has 991,600 people living with HIV and has expanded access to annual HIV viral load (VL) testing to enhance care quality for clients. However, significant delays persist in returning VL results from laboratories to facilities and from sample collection to client receipt. To address this, we implemented a digital viral load results return (VLRR) application, using existing mobile phone platforms (SMS/USSD), to expedite results return to clients and healthcare providers (HCPs).

**Methods:** VLRR is a digital SMS/USSD platform leveraging mobile phones to reduce turnaround time (TAT) and improve access to VL results. To evaluate the VLRR intervention, we: (1) estimated the TAT for digital results return and compared it to the paper-based process, (2) calculated open rates for digital results, (3) conducted a qualitative evaluation with VLRR users, and (4) estimated the potential cost savings from avoiding unnecessary sample redraws.

**Results:** From April 2022 to June 2024, HCPs registered 4,067 clients. Before VLRR implementation, the average TAT for clients to receive VL results was 128 days. After VLRR enrollment, it was 48.5 days (a 62.4% improvement) using electronic medical record data. By Q2 2023, VLRR clients and HCPs received results in an average of 30 and 38 days.

The overall open rate for digital results (opened by either a client or HCP) was 72%. Of the clients interviewed, 98% expressed a desire to continue using VLRR, and 100% of HCPs recommended expanding the platform to other HCPs.

**Conclusion:** VLRR is effective in reducing TAT and improving access to VL results for both clients and HCPs. To enhance uptake and achieve national scale, VLRR can be integrated into Malawi’s existing EMR systems, further reducing TAT and enabling HCPs to deliver higher quality care and improve long-term clinical outcomes.

## Introduction

Timely turnaround times (TAT) for diagnostic results from laboratories to healthcare providers (HCPs) and clients are essential for quality care.^1-3^ Delays and backlogs in laboratory results hinder timely diagnosis and treatment.^4^ Excessive backlogs can render results clinically irrelevant, requiring blood sample redraws and restarting the process. This increases laboratory workload and strains the healthcare system both in time and financial resources.^5^ Faster TAT is crucial for effective client management.^4^

HIV is the leading cause of death and disability in Malawi, with approximately 991,600 people living with HIV (PLHIV), representing 4.6% of the population.^6^ Since the World Health Organization (WHO) released antiretroviral therapy (ART) guidelines in 2013, viral load (VL) testing has been the gold standard for monitoring ART adherence.^7^ Malawi’s standard of care recommends a VL test six months after ART initiation and, if virally suppressed, every year thereafter, with results delivered to facilities within 21 days and to clients within 30 days.^8^ However, Malawi has only 14 specialized laboratories to process VL samples for clients across 850+ ART facilities, leading to long processing times, laboratory backlogs, and extended travel times to clinics. As a result, these standards are rarely met, impacting the quality of care for PLHIV and long-term client outcomes. To meet the Joint Programme on HIV/AIDS (UNAIDS) treatment target of greater than 95% viral suppression among PLHIV on ART^9^, Malawi must innovate its laboratory information systems and significantly reduce TAT for VL testing.

There is evidence that providing laboratory results directly to clients via mobile phone technologies can reduce negative clinical interactions and mitigate related consequences. Further to that, Malawi sought to amplify the undetectable=untransmittable (U=U) campaign by implementing a viral load results return system that directly notifies clients of their undetectable status, empowering them with knowledge and promoting HIV prevention. This approach also empowers clients to seek the services they need to manage their condition. Zambia and Zimbabwe have piloted SMS interventions that sent results to facilities and reminders to clients to visit the facility for their results.^10-12^ In South Africa, the National Health Laboratory Services piloted a mobile platform called iThemba, a smartphone application that delivered HIV viral load results, education, and clinical support directly to users’ smartphones.^13^

These interventions demonstrated that the use of mobile phone technologies to deliver results to clients can improve the client experience. Providing clients with their results via mobile phone can limit the potential for negative encounters, provide positive encouragement, reduce delay in knowledge about one’s health status, and increase health-seeking behavior, which has a direct effect on longevity and the quality of a client’s life.^14^ We set out to determine the feasibility and acceptability of sending VL results to healthcare providers and clients using a combination of SMS and USSD. To our knowledge, no country has leveraged both SMS and USSD-based technology to transmit results. Subsequently, most countries did not report an open rate of the SMS or application. Similarly, no applications targeted the system-wide benefit that a mobile phone results return platform could have on the overall efficiency of the laboratory system.

We hypothesized that a cost-effective SMS/USSD platform that relayed confidential and timely results return could be created for all phone users (feature and smartphone) in Malawi to reduce TAT. To test this hypothesis, we developed a digital SMS and USSD-based platform to deliver VL results directly to clients and HCPs to reduce TAT and improve access to VL results with a high open rate. We evaluated this in four ways: 1) by estimating the TAT, 2) by estimating the result open rate, 3) through a formal qualitative evaluation of the clients and HCPs using the application, and 4) by evaluating the system-wide benefit of the application by estimating the number of redraws that could be offset through near-real-time results return.

## Methodology

The Viral Load Results Return (VLRR) application was launched as a pilot in February 2022 in four health facilities across Malawi. VLRR was extended to 5 additional sites in October 2023 and an additional 5 sites in January 2024. We used a mixed methods approach to evaluate the platform’s success at all 14 health facilities across 13 districts.

We selected study sites based on the 3 regions of Malawi, rural/urban status, ART cohort size (>900 clients), affiliation (public vs private/semi-private), and access to electronic medical records. We enrolled clients who were above 18 years old, due for a routine VL test, and consented to receive their VL result through a mobile phone at the selected study sites.

We used quantitative methods to determine the feasibility of the intervention to effectively return results by estimating the TAT for both clients and HCPs. We conducted a comparative analysis between the proportion of clients accessing results using VLRR compared to the paper-based system (e.g., result open rates). We used qualitative methods to determine the acceptability of the platform through in-depth interviews with both clients and healthcare providers. The in-depth interviews with healthcare providers and clients gauged their perception of the platform’s ease of use, accessibility, user support, privacy, and security.

Lastly, we conducted a cost analysis of the sample redraws required due to long TAT.

### SMS/USSD Application Intervention

The VLRR application is a digital SMS and USSD-based platform that leverages mobile phones (both feature and smartphone) to deliver results directly to clients. HCPs at facilities who provided HIV services were also notified via SMS that clients’ results were ready, and they could also view the results. This was intended to reduce the TAT and improve access to VL results by circumnavigating some of the challenges within the paper-based system (e.g., lost results, lab backlogs, result delivery times, client travel time). We leveraged a combined SMS and USSD platform to make it accessible for non-smartphone users. We used SMS to send the notifications that a result is ready and USSD to privately check results. Results can only be viewed within USSD to protect client privacy and confidentiality, as USSD times out after 30 seconds.

We used routine Laboratory Information Management System (LIMS) and VLRR data for these analyses. The LIMS database houses routine VL results from all 14 laboratories in Malawi. The VLRR platform’s Application Programming Interface (API) is connected to the LIMS database. When a laboratory approves a result in LIMS, it is synched to VLRR which triggers a nondescript SMS to be sent directly to a client’s phone notifying them their result is ready. The SMS prompts the client to dial the *929# shortcode to check their VL result in the USSD platform. The *929# shortcode was granted to the Ministry of Health (MOH) by the Malawi Communications Regulatory Authority (MACRA). It was initially utilized for COVID-19 and was extended to other digital services, including VLRR. Additionally, the MOH supported the process of connecting VLRR to the two most used telecommunication companies in Malawi. Figure 1 shows the VLRR workflow for clients. Additionally, HCPs who were trained on and use the VLRR platform receive an SMS, Monday through Friday, with a list of samples that have results ready. An HCP checks a client’s result within the USSD platform using the client’s ART number. If the VL result is high, the HCP is prompted to send out a community health worker or an expert client to retrieve the client in the community and bring them back to the clinic.

**Figure 1.**
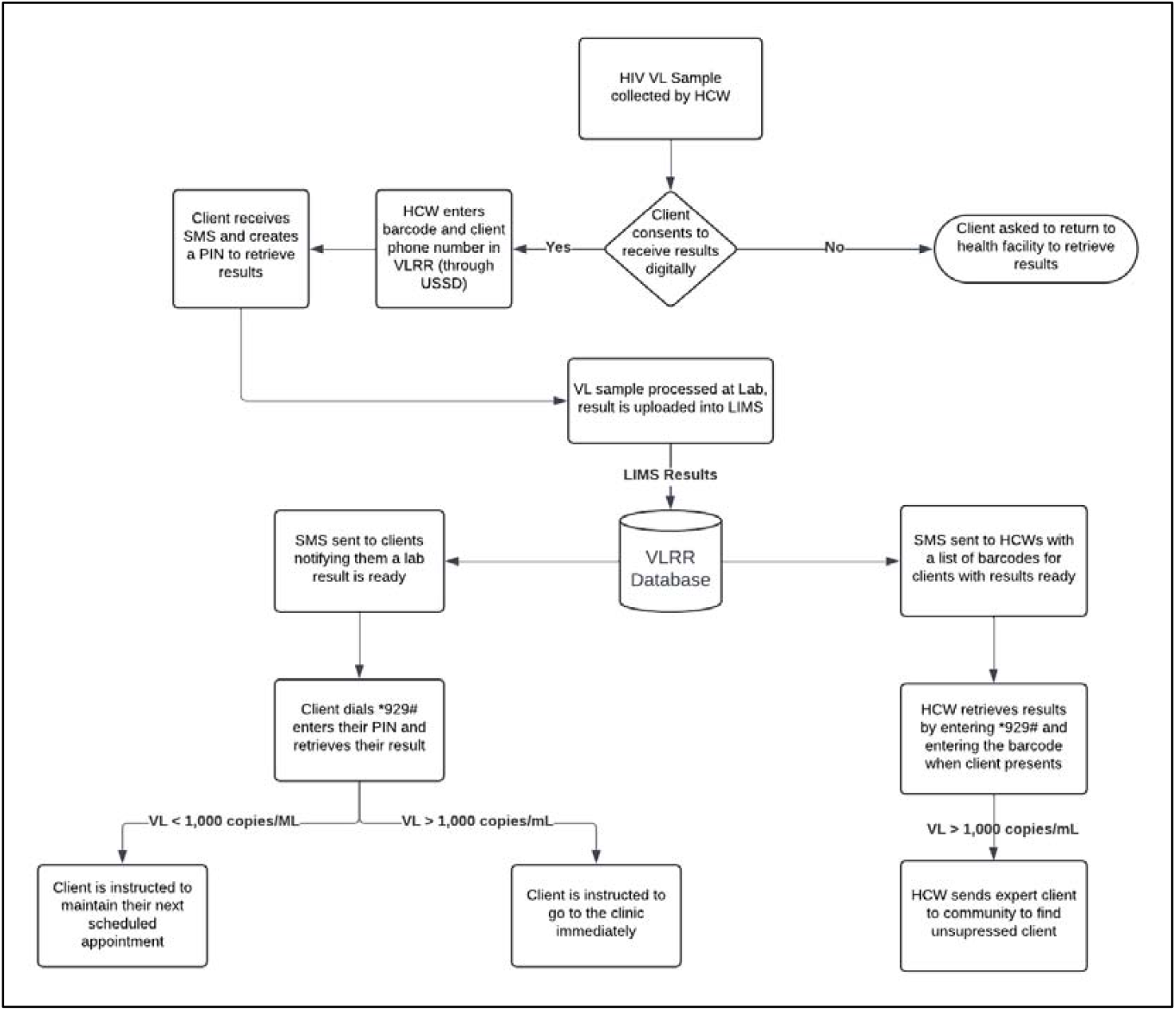
VLRR Workflow for clients and HCPs.

#### 1) Turnaround Time

When a client on ART gets a VL sample taken at the facility, the HCP asks them if they want to receive their results digitally. If the client consents, the HCP enrolls them into the VLRR platform, helps the client set up their personal identification number (PIN) in the USSD platform, and teaches the client how to navigate the USSD platform. The client learns how to dial the USSD shortcode, enter their PIN, and check their VL results through this process.

We used TAT and open rates to determine the successful delivery of results by the platform to clients and HCPs. We analyzed routine data collected by the platform throughout the pilot period. We calculated TAT from the moment a client had a sample drawn to when an SMS was sent to both the client and the HCPs. We also calculated TAT from sample draw to when a result was opened by a client and a HCP. We monitored and calculated TATs throughout the lifecycle of the intervention and compared the percent change between the status-quo TAT (pre-VLRR) and VLRR’s TAT. To estimate the status quo national TAT, we used electronic medical record (EMR) data from 249 EMR sites from January 2019 to May 2024. We also subset EMR data to the 11 VLRR EMR sites to estimate the status quo TAT for VLRR sites only. Three VLRR sites are non-EMR-enabled sites.

#### 2) Application Open Rates

To determine open rates, we tracked the percentage of clients and HCPs who were sent an SMS notifying them that a result was ready. We calculated open rates for VLRR as the percentage of clients or HCPs who viewed the result on USSD after getting the SMS notification. We compared the open rates for VLRR with those of existing results return digital platforms in sub-Saharan Africa and high-income countries.

#### 3) Qualitative Evaluation of the Application

We engaged four Malawian research assistants (RAs) to conduct qualitative interviews with clients and HCPs.

They interviewed HCPs who were trained on and used the VLRR platform. We aimed to interview 5 healthcare providers per site. However, some sites contributed lower numbers as some providers were away on leave or assigned other tasks, thereby limiting their availability for interviews. We interviewed a total of 67 healthcare providers. Table 1 shows the descriptive statistics for HCPs included in the qualitative study including sex breakdown, average age, average time working in HIV, and average period using the platform.

We aimed to interview 10 clients per health facility. However, some facilities contributed fewer numbers of clients. Reasons for the fewer numbers included a lack of interest to participate in the interviews, transportation challenges in certain areas due to flooded rivers, and prioritization of farming activities over coming to the facility for interviews. We obtained clients via purposive sampling. HCPs reached out to clients who were enrolled in the VLRR platform and asked them of their interest to participate in the interviews. We interviewed a total of 117 clients. Table 2 shows the descriptive statistics of the clients interviewed across the 14 sites.

Before each interview, the RAs obtained written consent from all study participants. If a participant could not read or write, the RAs read out the consent form to the participant and obtained written consent using a thumbprint. The RAs conducted the interviews using questionnaires and voice recorders. The questionnaires for the clients and HCPs were different and consisted of a combination of qualitative and Likert scale questions. All questionnaires were verbally administered to healthcare providers in English, while clients were provided the option of being interviewed in either English or Chichewa. All interviews conducted in Chichewa were recorded, translated, and transcribed into English.

For clients, the survey evaluated clients’ favorability of the platform and whether they would recommend VLRR to others living with HIV, ease of accessing the results and overall perceptions of VLRR, and any privacy and security concerns. For HCPs, we evaluated how they felt VLRR supported clients, the challenges they faced with the paper-based process and whether VLRR addressed those challenges, ease of use of the platform, VLRR’s effect on their overall workload, and continuity of VLRR.

We thematically coded all qualitative questions and categorized them based on responses. These categories were not mutually exclusive. Therefore, a single respondent could have an answer that fits into several categories.

#### 4) Cost Analysis - Redraws

To quantify the potential cost savings from unnecessary sample redraws, we used LIMS data to calculate the total number of samples taken in a year that were redrawn due to clinical irrelevance. Clinically irrelevant VL result is defined as a result delivered to the facility after 3 months from sample collection since it can no longer be used to inform the care of the client. It is necessary to note that current data systems in Malawi, including LIMS and the Electronic Medical Records (EMR), only track redraws due to laboratory error. These systems do not capture redraws necessitated due to clinical irrelevance.

To estimate the number of tests that required a redraw due to clinical irrelevance, we calculated the proportion of clients with a low VL who had two tests within 5 months using EMR data for the 11 EMR-enabled sites. We chose this period because the Malawi Integrated Guidelines for Clinical Management of HIV 2022^15^ indicate that a client with a low VL should receive a VL test no earlier than 6 months after their last low VL test.

We then estimated the all-in cost per VL laboratory test from the lowest to the highest possible costs. Based on an assessment conducted by the Médecins Sans Frontières^16^, the all-in (e.g., test, labor, time, transport, etc.) cost to conduct a VL test is ∼$35·38 site MSF’s reagent costs in Malawi. We also estimated the commodity cost of a viral load test to be ∼$11.50 USD as a minimum, provided by the Malawi Ministry of Health Laboratory Specialist.

We multiplied the estimated number of VL sample redraws against the estimated cost per test to determine the potential cost savings that VLRR would introduce by returning the results within the clinically relevant period for the 11 EMR-enabled sites. We then performed this same analysis and predicted the potential cost savings if VLRR were to be scaled at the national level.

### Role of the funding source

The Gates Foundation funded and supported this work, but did not participate in the study design, data collection, analysis, interpretation, or decision to publish these results.

## Results

We have categorized our findings based on TAT, open rates, and qualitative feedback from clients and HCPs. Site-specific details can be found in Table 1.

**Table 1.** Site Level VLRR Characteristics, Open Rates, and TATs.

| Pilot Phase | Rural/Urban | District | Site ID | Samples Registered | Results Sent | Results Opened by Clients | Results Opened by HCPs | EMR enabled |
| --- | --- | --- | --- | --- | --- | --- | --- | --- |
| Initial | Rural | Lilongwe | Site 1 | 528 | 479 (91%) | 78 (16%) | 269 (56%) | Yes |
| Initial | Rural | Mzimba | Site 2 | 117 | 111 (95%) | 14 (13%) | 89 (80%) | Yes |
| Initial | Rural | Machinga | Site 3 | 205 | 189 (92%) | 26 (14%) | 94 (50%) | Yes |
| Initial | Urban | Lilongwe | Site 4 | 788 | 689 (87%) | 175 (25%) | 242 (35%) | Yes |
| Scale-Up | Rural | Mulanje | Site 5 | 419 | 328 (78%) | 10 (3%) | 104 (32%) | Yes |
| Scale-Up | Rural | Kasungu | Site 6 | 112 | 78 (70%) | 23 (29%) | 59 (76%) | No |
| Scale-Up | Rural | Neno | Site 7 | 44 | 37 (84%) | 6 (16%) | 30 (81%) | No |
| Scale-Up | Rural | Salima | Site 8 | 70 | 62 (89%) | 12 (19%) | 53 (85%) | Yes |
| Scale-Up | Rural | Dedza | Site 9 | 259 | 230 (89%) | 20 (9%) | 47 (20%) | No |
| Scale-Up | Urban | Zomba | Site 10 | 489 | 464 (95%) | 228 (49%) | 195 (42%) | Yes |
| Scale-Up | Urban | Rumphi | Site 11 | 161 | 149 (93%) | 35 (23%) | 55 (37%) | Yes |
| Scale-Up | Urban | Chiradzulu | Site 12 | 226 | 207 (92%) | 73 (35%) | 149 (72%) | Yes |
| Scale-Up | Urban | Mchinji | Site 13 | 457 | 380 (83%) | 124 (33%) | 353 (93%) | Yes |
| Scale-Up | Urban | NkhataBay | Site 14 | 192 | 181 (94%) | 65 (36%) | 46 (25%) | Yes |
|  | Total |  |  | 4067 | 3584 (88%) | 889 (25%) | 1785 (50%) |  |

### 1) Turnaround Time

Over the life of the intervention, HCPs successfully registered 4,067 clients (Table 1). Of those registered clients, 88% received an SMS (Table 1). Clients did not receive an SMS due to initial data entry error of client details at initial registration. The average TAT in Malawi for the status-quo process using EMR data from 249 sites is 85·9 days (from sample collection to the client’s next return visit).

For VLRR, the average TAT for results return (sample collection to results receipt) at all 14 sites declined over time for both clients and HCPs. In the last quarter of 2023, the average TAT was 47 days for clients and 53 days for HCPs. By the end of Q2 2023, the average TAT reduced to 30 days for clients and 38 days for HCPs (Figure 2). The overall average TAT from Q3 2023 to Q2 2024 was 49 days for clients and 56 days for HCPs. We report from Q3 2023 to Q2 2024 because in Q2 2023, we resolved a back-end synchronization issue delaying results return as well as implemented an improved user workflow to streamline the process of checking results for clients.

This explains the steady decline and improvement in TAT results for both HCPs and clients as shown in Figure 2.

**Figure 2.**
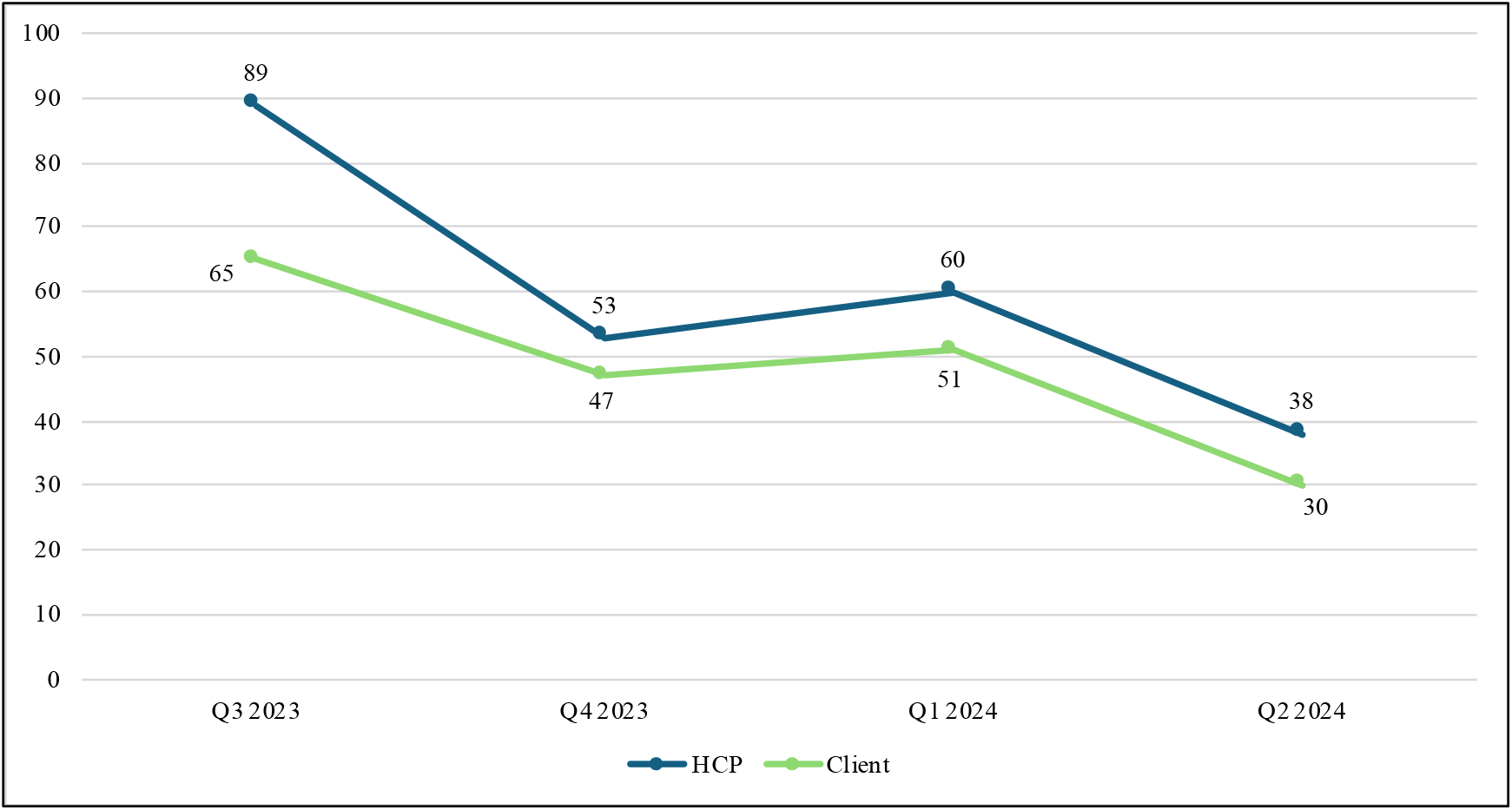
Quarterly TAT (days) for Clients and HCPs from Q3 2023 to Q2 2024 enrolled in VLRR.

For the 11 VLRR EMR sites, the client’s TAT (sample collection to next facility visit) pre-VLRR enrollment was 129 days from January 2019 to May 2024. After VLRR enrollment, these same clients’ TAT reduced to 48·5 days from Q3 2023 to Q2 2024 to view their results. This is a 62·4% improvement in TAT for those clients who enrolled in VLRR.

### 2) Application Open Rates

The overall open rate, or the rate a result was opened by either a client or HCP, was 72%. The individual open rate for clients and HCPs was 25% and 50%, respectively (Table 1). For clients, quarterly open rates stayed relatively consistent from Q3 2023 to Q2 2024 with only slight increases in Q4 and Q1 2024 (Figure 3).

However, HCPs had more variability in their open rate compared to clients over the same period with a big jump from Q3 2023 to Q4 2023 (Figure 3), which then slightly declined over time. This could be explained by a more hands-on approach with HCPs from the study team. Throughout the project, the study team along with DHA officials held quarterly site supervisions to engage with HCPs and identify if they were having any issues with the platform. The study team did not interact with clients directly throughout the life course of the intervention.

**Figure 3.**
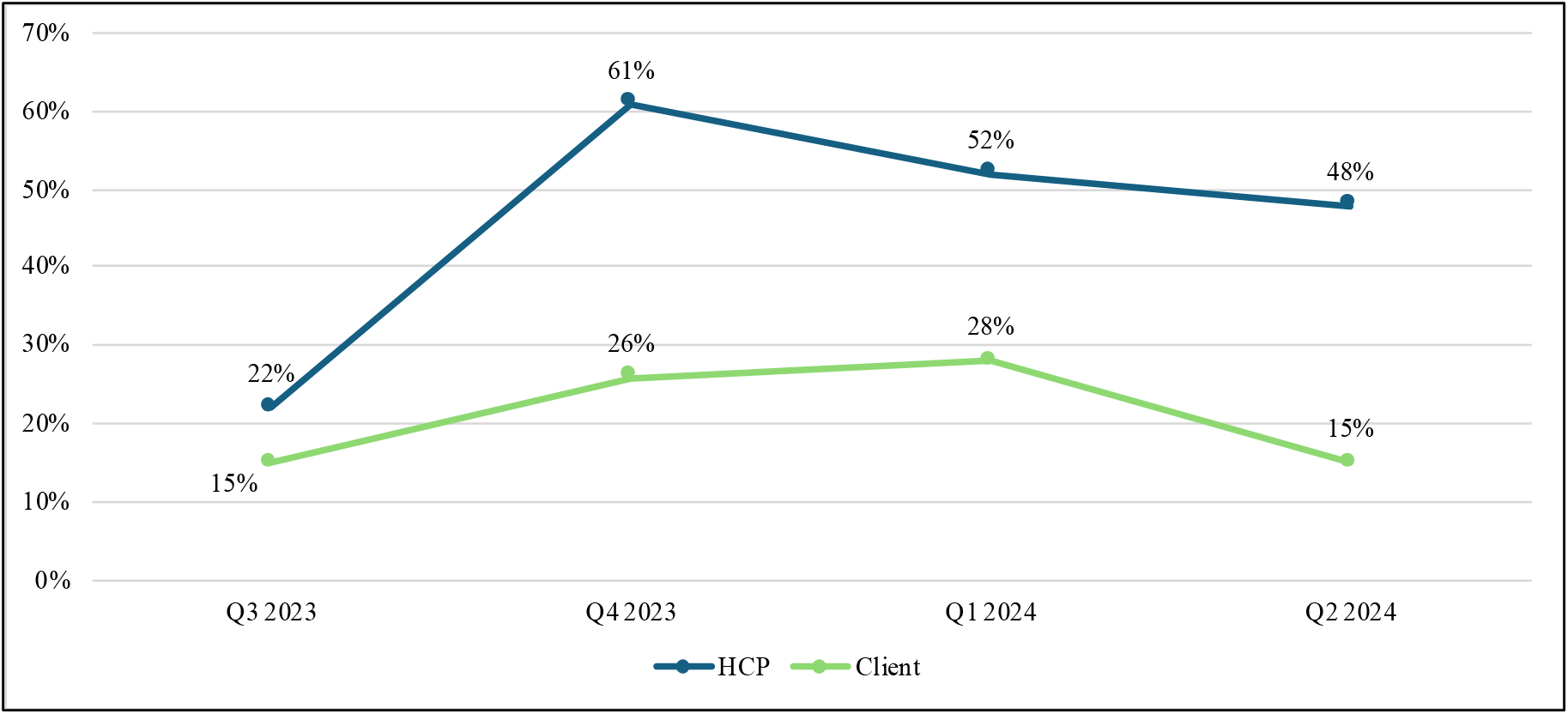
Quarterly Open Rates for Clients and HCPs from Q3 2023 to Q2 2024.

### 3) Qualitative Evaluation

The findings from the qualitative evaluation support the quantitative findings of decreased TAT and improved open/access rate to results.

#### Client Results

We interviewed 117 clients across the 14 facilities. Of these, there were slightly more females than males (67 vs 50). The average age for clients interviewed was 43 years, with a range of 18-76 years. The average length of time a client was on ART was ∼12 years, and the average length of VLRR enrollment was 4 months (Table 2). All clients, except one, indicated they had access to a phone. When asked how long clients typically waited for their paper VL results, the average wait time was 15 weeks across facilities, but some waited up to 48 weeks to receive their paper results (Table 2).

Overall, 98% of clients liked the platform and would recommend it to others who need to receive a VL result. Further, 86% of clients felt they received enough support from HCPs to access and use the platform adequately.

When a client was asked a free-form response about the benefits of accessing their results through the VLRR platform, 38% (Table 2) of clients indicated they liked that it saved them time and money by avoiding long travel distances to facilities and not having to stop work to go get their results. This is a high level of agreement seen in an open-ended qualitative response. Similarly, 7% (Table 2) liked that they avoided long clinic visits/additional clinic visits, and 12% of clients expressed favor in being able to access their results from anywhere, especially at home (Table 2). Over 30% (Table 2) of clients were surprised that they could receive their results on time/quickly and felt it gave them the ability to take proper action based on their results. Other clients expressed a sense of agency in their care, getting to know their results on their own and being able to go to their provider already knowing their results.

We also sought to understand clients’ perceptions of VLRR and privacy. While most clients had no concerns about privacy (96%) and indeed expressed that VLRR increased their privacy, 4% expressed a level of concern surrounding their privacy and VLRR, especially if someone used their phone. However, many caveated this concern, by noting that the PIN eased their concerns since no one else knew their PIN and could access their results.

Clients overwhelmingly liked and wanted to keep using the platform (98%) and said the platform was easy to use (91%). Additionally, they wanted their friends and family to have access to it (98%). Clients preferred VLRR because it saved them time by eliminating the need to travel to the clinic, saved them money by avoiding missed work, and provided them with fast results (56%). They also got excited about knowing their results before visiting the clinic and expressed it gave them more agency to act if a clinical change was necessary.

**Table 2.**
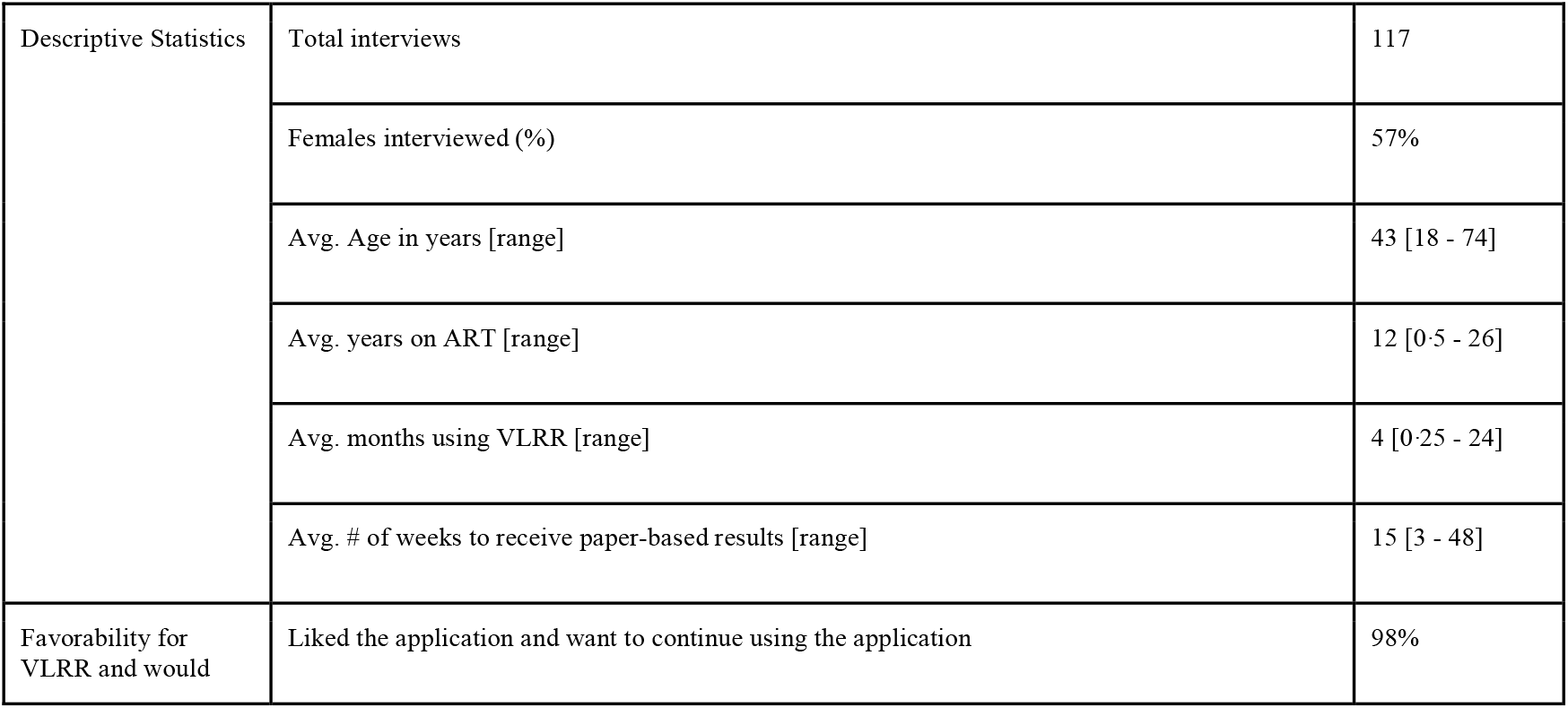
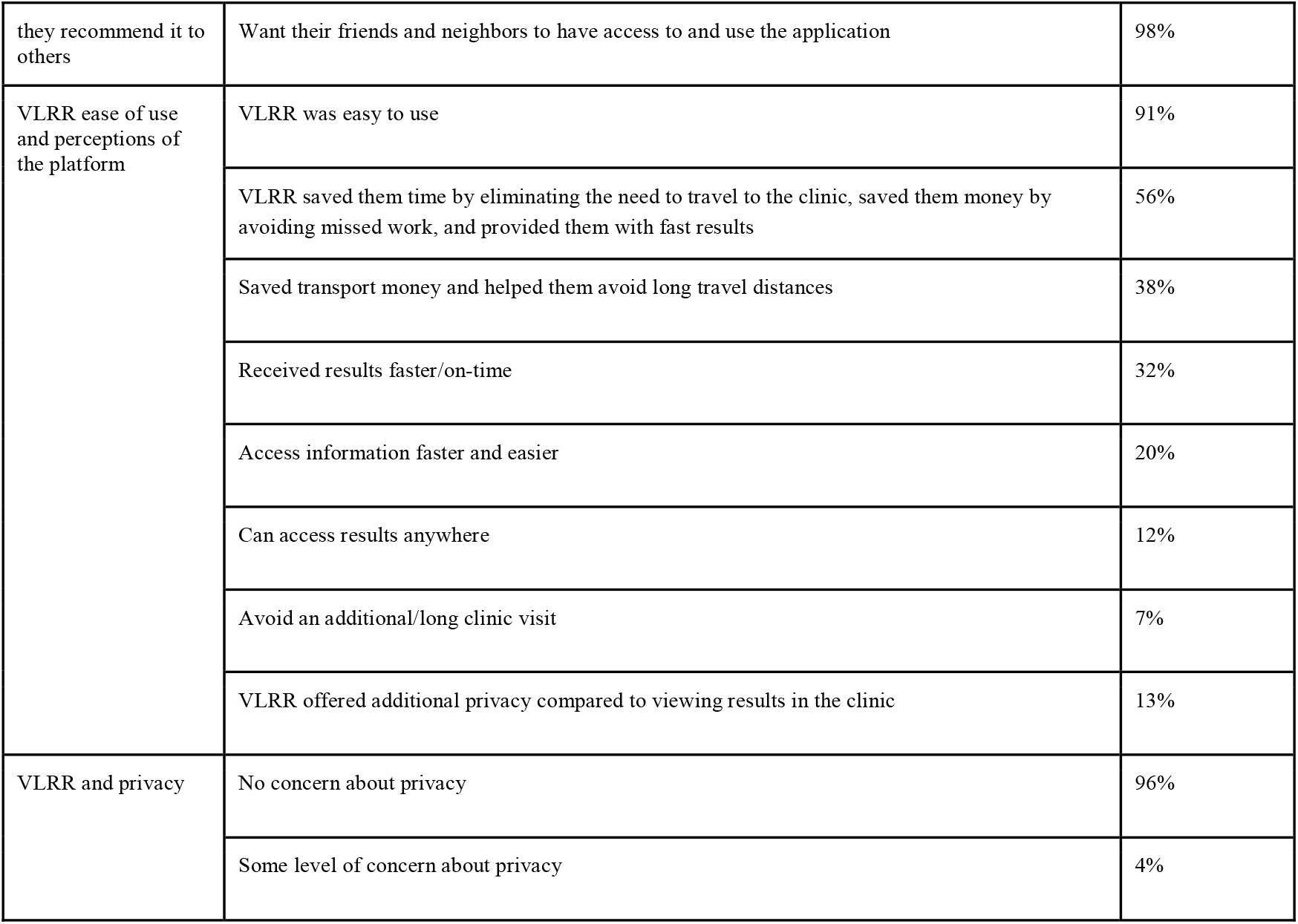
Client results from the qualitative evaluation for all 14 facilities HCP Results.

#### HCP Results

We conducted interviews with 67 HCPs across the 14 facilities. The sample included 35 males and 32 females. All HCPs in our sample were a part of the VLRR training and had registered clients (Table 3).

The average age of HCPs interviewed was 39 years, with a range of 23-59 years. The average amount of time HCPs had worked in HIV care was about 9 years and the average length of time using VLRR was 7 months (Table 3). The most common cadres represented were nurses (18%) and health diagnostic assistants (27%). About 50% of HCPs had some level of college education.

Overall, HCPs supported the VLRR application. Ninety-seven percent (97%) of HCPs were likely/very likely to recommend the application to other HCPs and 97% of HCPs thought the VLRR application improved client access to results.

When reflecting on what made VLRR successful through free-form responses, HCPs stated that the VLRR application supported clients to receive their results quickly and easily (54%) and allowed them to access results anywhere, especially at home (27%) (Table 3). HCPs also said it supported clients receiving results because they had to do less tracing since clients came back earlier than their next scheduled appointment, if unsuppressed (9%) (Table 3).

Notable challenges of the paper-based system for healthcare providers included long TAT (72%), long clinic visits/travel time (9%), and if there would be an error with the results, they would need to recollect the sample (3%) (Table 3).

When asked in an open-form question if they felt the VLRR platform addressed the challenges of the paper-based process, nearly half of HCPs said yes, it reduced TAT. Further, 12% said it reduced client travel time, and 12% noted that it increased patient action and knowledge of results before their next scheduled appointment (Table 3). Additionally, 76% of HCPs strongly agreed that it was easy to enter client information and 89% found results easy to access through VLRR.

Over 20% of HCPs said VLRR reduced/simplified their workload by reducing the number of clients coming to the facility to get their results, and 12% said it enhanced their work quality (Table 3). Many HCPs noted their favorability for the platform because clients knew their results, which helped them to build trust with their clients by enhancing the reliability of clients receiving their results on time.

Additionally, 100% of HCPs said the platform should continue and should be scaled to additional facilities. We asked them why or why not they would want VLRR to continue and 27% said it reduced TAT for results, 19% said it reduced workload, and 21% said overall, it was a good initiative, and the technology was useful. Further, HCPs noted that with the paper-based process, clients often would come back to the facility and their results would not be ready. This reduced client trust in the facility as well as reduced their desire to redraw their test and ensure they were truly virally suppressed, thereby affecting clinical care and outcomes.

**Table 3.** HCP results from the qualitative evaluation for all 14 facilities.

|  |  |  |
| --- | --- | --- |
| Descriptive Statistics | Total Interviewed (#) | 67 |
|  | Females Interviewed (%) | 48% |
|  | Avg. Age in years [range] | 39 [23 - 59] |
|  | Avg. years working in HIV [range] | 9 [0.25 - 20] |
|  | Avg. months using VLRR [range] | 7 [1 - 24] |
| How VLRR supported clients | Thought the VLRR application improved client access to results | 97% |
|  | VLRR supported clients to receive their results quickly and easily | 54% |
|  | VLRR allowed clients to access results anywhere, especially at home | 27% |
|  | VLRR supported clients receiving results because they had to do less tracing since clients came back earlier than their next scheduled appointment, if unsuppressed. | 7% |
| Challenges HCPs faced with the paper-based results return process and did VLRR address those challenges | The paper-based process has a long TAT | 72% |
|  | Clients experience long clinic visits/travel time with the paper-based process | 9% |
|  | There are errors with the paper results and the client must come back in for a redraw | 3% |
|  | VLRR enhanced their work quality because clients knew their results when they would come in for a clinic visit. This helped them to build trust with their clients by enhancing the reliability of clients receiving their results on time. | 12% |
| Ease of use | It was easy to enter client information when registering clients in the platform | 76% |
|  | Results were easy to access through the USSD platform | 89% |
| VLRR and HCP workload | Were likely/very likely to recommend the application to other HCPs | 97% |
|  | VLRR reduced/simplified their workload by reducing the number of clients coming to the facility to get their results | 20% |
| Should VLRR continue and why | VLRR should continue and should be scaled to additional facilities. | 100% |
|  | Why should VLRR continue: Reduces long TAT | 27% |
|  | Why should VLRR continue: Reduces workload | 19% |
|  | Why should VLRR continue: Increased client trust | 12% |

### 4) Cost Analysis - Redraws

The VLRR intervention also has a future opportunity to provide system wide benefits and cost savings by reducing the overall laboratory backlog by reducing unnecessary redraw tests due to long TAT. In Malawi, there are approximately 50,340 viral tests drawn each year at the 11 EMR-enabled VLRR sites. Approximately 30% (15,100) of those tests were avoidable redraws taken due to long TATs, which led to clinically irrelevant VL results. This 30% accounts for virally suppressed clients who had two tests within a 5-month period. If this 30% was avoided through expedited turnaround time of results, this would result in a potential cost savings of $173,650 to $534,238.

At the national level, Malawi draws about 675,000 VL tests per year. Of those 675,000 approximately 25-28% of those are avoidable redraws. Using the MSF comprehensive cost of $35·38, savings would be $6·7 million. Using the reagent cost of $11,·50, the minimal cost savings would be $1·8 million.

## Discussion

Adequate clinical decision-making relies on the timely return of laboratory results.^1-3^ Delayed results inhibit clinical care, increase the cost and burden to HCPs and the healthcare system, and deteriorate the trust between provider and client.^5^ WHO emphasized the importance of VL monitoring in their 2013 guidelines for improving clinical outcomes^7^; however, adequate VL monitoring is not feasible if results are lost, delayed, or required to be redrawn. The VLRR intervention is a simple and cost-effective digital health intervention that leverages existing government structures and existing back-end architecture in Malawi to improve TATs of laboratory results and increase access to results for both clients and HCPs faster than the paper-based process.

Prior to VLRR, Malawi had increased access to annual HIV viral load testing to improve the quality of care for clients. However, the VL results were still returned to clients in person, leading to delays, lost results, and missed opportunities for timely clinical interventions. This had significant implications for unsuppressed clients requiring adherence counseling or regimen changes and created additional backlogs at laboratories. VLRR addressed these issues by significantly reducing the turnaround times for VL results and ensuring both HCPs and clients could access their results more quickly and reliably from any location.

In addition to decreasing TAT, this intervention also intended to create a user-friendly digital solution that would increase client and HCP access to results, which we assessed through open rates. We consider the overall open rate to be more clinically relevant as action will be taken by *either* the client or HCP if needed. For example, if a client opens their results and finds they are unsuppressed, they are guided to visit the clinic as soon as possible. Similarly, if a HCP opens a client’s result and finds a client to be unsuppressed, they will send an expert client or community health worker to bring the client back to the facility to receive enhanced adherence counseling (EAC) or be switched to 2nd line therapy, as appropriate.

VLRR is the first application to leverage a combined SMS/USSD results return platform to clients in low- and middle-income countries. Because VLRR is the first intervention of its kind, there are no directly comparable open rates to measure success. However, we sought to understand open rates to digital applications providing test results to clients in both low and high-income countries^14, 17-25^ and assess how VLRR compared to these. The open rates for high-income countries ranged from 21% to 57%. The VLRR overall open rate was about 15% to 51% higher, compared to the best^23^ and least^21^ performing applications in these countries, respectively. For low-income countries, most interventions did not report an open rate, except for one smartphone application, which had a high viewership rate of 78%^23^. While not directly comparable, VLRR’s overall open rate (72%) performs better or on par with reported smartphone and SMS interventions.

The faster TAT and usage rate of the VLRR platform are supported by the qualitative findings, which demonstrate the overall satisfaction that clients and healthcare providers have with the VLRR platform. Clients and healthcare providers alike appreciate how fast results are returned to them and the ease with which they can access them. One of the main objectives of VLRR was to enhance the speed at which clients and HCPs accessed viral load results. The qualitative evaluation confirms that both clients and HCPs felt a palpable difference in TAT, liked the ease with which they could access results, and the benefits of accessing results at any time from any location, in private.

Further, the VLRR digital solution demonstrates the potential for system-wide benefits and essential cost savings by offsetting the overall laboratory backlog by targeting unnecessary redraw tests necessitated by long TATs. Nationally, this equates to $1·8-$6·7 million in potential cost savings. The cost savings have implications for the government, districts, facilities, and ultimately, clients.

The VLRR platform was designed with broadly accessible technology and adaptable architecture, allowing for seamless expansion to accommodate other functions. The VLRR platform can be leveraged for additional use cases including other tests (e.g., TB, malaria, HPV), appointment reminders, precision nudging to recheck results, and an AI chatbot for smartphone users. These use cases could be added to the platform with minor tweaks to the backend architecture and provide opportunities to lower average care costs by improving efficiency and reducing the excess burden of disease through enhanced clinical continuity. The platform extensions leverage the existing VLRR frontend and backend architecture. These additional use cases can increase access to lab results and improve the standard of care for clients using this simple and cost-effective technology.

### Limitations and Lessons Learned

The VLRR TAT was variable to many exogenous factors that could not be controlled by the study team. Some of these factors included telecommunication company service disruptions, nationwide power outages, client literacy rates, clients changing SIM cards, commodity stockouts, and server capacity. During periods of downtime, we would notify all HCPs via WhatsApp that VLRR was down and would be back up as soon as possible and communicated as best we could on how the issue would be resolved.

To address challenges around literacy, we tried to simplify the process by implementing the PIN workflow for clients. This would reduce the amount of information they would have to enter to access their results, but still respect their privacy and maintain confidentiality because the PIN they chose was specific to them.

We also experienced data quality challenges with EMR data and incorrect data entry of ART numbers and sample barcodes. We matched as many samples as possible based on multiple client identifiers. However, this, at times, created challenges to calculate the most accurate TAT. Additionally, non-EMR sites do not have a baseline TAT besides past studies that have estimated TAT. So, it becomes a challenge to compare VLRR’s TAT to an existing metric for Malawi. However, we utilized the data provided by the specimen transport service to estimate the TAT for non-EMR sites to have a rough estimate of comparison.

## Conclusion

Despite the challenges faced during the VLRR intervention, the application has achieved many more successes for both HCPs and clients accessing viral load results faster than the status quo paper-based processes. The VLRR application demonstrates that long TATs can be mitigated with simple digital solutions. VLRR has demonstrated the improvements that can be made by targeting gaps in the process of sample collection to results return. To further enhance uptake and national scale of the platform, VLRR can be integrated into existing point-of-care systems within Malawi (EMR) to continue to reduce TAT and support providers to treat clients adequately and timely, to continue improving clinical outcomes long-term. Further, this technology can be easily adapted and leveraged in other countries to support faster results return for HIV-positive clients to support the improvement of global suppression rates.

## Acknowledgements

Not applicable.

## Author Contributions

RH is the corresponding author and led survey design, data management, data analysis, and manuscript writing. CM supported survey design, data collection, data analysis, and manuscript review. BK supported the design of the work, data analysis, and manuscript review. LM supported data analysis and manuscript review. TS supported the design of the work and manuscript review. HC supported the design of the work and manuscript review. BC supported district and facility coordination as well as manuscript review. LC support project supervision at the district level and manuscript review. DG supported data analysis, project conceptualization, and manuscript review.

## Statements and Declarations

Not applicable.

## Ethical Considerations

Ethical clearance was approved and obtained by the National Health Research Ethics Committee in Malawi for the digital intervention as well as conducting the formal qualitative evaluation.

## Consent to Participate

Upon registration, all VLRR users digitally consented to use the platform and receive SMSs. Additionally, all participants in the qualitative evaluation provided written consent to participate and could opt out of the interview at any time.

## Consent for Publication

Not applicable.

## Declaration of Conflicting Interest

The author(s) declared no potential conflicts of interest with respect to the research, authorship, and/or publication of this article.

## Funding Statement

Funding for this project was provided by the Gates Foundation.

## Data Availability

Aggregate SMS/USSD data can be shared upon request. Other data cannot be shared as it is personally identifiable health information.

## Notes

### Competing Interest Statement

The authors have declared no competing interest.

### Funding Statement

This study was funded by the Gates Foundation.

### Author Declarations

Ethical approval was granted by the National Health Science Research Ethics Committee in Malawi. Additionally, this study was overseen and approved by the Department of HIV/AIDS, Malawi Ministry of Health.

## References

1. Erasmus RT, Zemlin AE. Clinical audit in the laboratory. J Clin Pathol. 2009;62(7):593–7. 10.1136/jcp.2008.056929

2. Hawkins RC. Laboratory turnaround time. Clin Biochem Rev. 2007;28(4):179–94. https://pubmed.ncbi.nlm.nih.gov/18392122/

3. Howanitz JH, Howanitz PJ. Laboratory results. Timeliness as a quality attribute and strategy. Am J Clin Pathol. 2001;116(3):311–5. 10.1309/H0DY-6VTW-NB36-U3L6

4. Lalongo C, Porzio O, Giambini I, Bernardini S. Total Automation for the Core Laboratory: Improving the Turnaround Time Helps to Reduce the Volume of Ordered STAT Tests. J Lab Autom. 2016;21(3):451–8. 10.1177/2211068215581488

5. Imoh LC, Mutale M, Parker CT, Erasmus RT, Zemlin AE. Laboratory-based clinical audit as a tool for continual improvement: an example from CSF chemistry turnaround time audit in a South-African teaching hospital. Biochem Med (Zagreb). 2016;26:194–201. 10.11613/BM.2016.021

6. UNAIDS. Malawi. Malawi Country Profile [Internet]. 2024 Jul 8. Available from: https://www.unaids.org/en/regionscountries/countries/malawi

7. World Health Organization. Consolidated Guidelines on the Use of Antiretroviral Drugs for Treating and Preventing HIV Infection: Recommendations for a Public Health Approach, 2nd ed. Geneva: WHO; [no date]. Available from: https://www.who.int/publications/i/item/9789241549684

8. LABORATORY AFRICAN HEALTH PROFESSIONS REGIONAL COLLABORATIVE. Scale Up of Viral Load Testing and Patient Management Project Proposal [Internet]. Available from: https://www.commonwealthnurses.org/ARC/Documents/Malawiproposal.pdf

9. UNAIDS. UNAIDS welcomes new research on “opt-out” HIV testing in England [Internet]. 2023 Nov 29. Available from: https://www.unaids.org/en/resources/presscentre/pressreleaseandstatementarchive/2023/november/20231129_new-research-on-opt-out-hiv-testing-england

10. Saito S, Duong YT, Metz M, Lee K, Patel H, Sleeman K, et al. Returning HIV-1 viral load results to participant-selected health facilities in national Population-based HIV Impact Assessment (PHIA) household surveys in three sub-Saharan African Countries, 2015 to 2016. J Int AIDS Soc. 2017;20 Suppl 7:e25004. 10.1002/jia2.25004

11. Seidenberg P, Nicholson S, Schaefer M, Semrau K, Bweupe M, Masese N, et al. Early infant diagnosis of HIV infection in Zambia through mobile phone texting of blood test results. Bull World Health Organ. 2012;90(5):348–56. 10.2471/BLT.11.100032

12. Venables E, Ndlovu Z, Munyaradzi D, Martínez-Pérez G, Mbofana E, Nyika P, et al. Patient and health-care worker experiences of an HIV viral load intervention using SMS: A qualitative study. PLoS One. 2019;14(4):e0215236. 10.1371/journal.pone.0215236

13. Lalla-Edward ST, Mashabane N, Stewart-Isherwood L, Scott L, Fyvie K, Duncan D, et al. Implementation of an mHealth App to Promote Engagement During HIV Care and Viral Load Suppression in Johannesburg, South Africa (iThemba Life): Pilot Technical Feasibility and Acceptability Study. JMIR Form Res. 2022;6(2):e26033. 10.2196/26033

14. Agarwal AK, Ali ZS, Shofer F, Xiong R, Hemmons J, Spencer E, et al. Testing digital methods of patient-reported outcomes data collection: Prospective Cluster Randomized Trial to test SMS text messaging and mobile surveys. JMIR Form Res. 2022;6(3). 10.2196/31894

15. Ministry of Health and Population, Malawi. Clinical Management of HIV in Children and Adults [Internet]. 2022. Available from: www.hiv.health.gov.mw

16. Médecins Sans Frontières Access Campaign. How low can we go? - pricing for HIV viral load testing in low- and middle-income countries [Internet]. Available from: https://msfaccess.org/sites/default/files/How%20Low%20Can%20We%20Go%20VL%20pricing%20brief.pdf

17. Oest SER, Hightower M, Krasowski MD. Activation and Utilization of an Electronic Health Record Patient Portal at an Academic Medical Center—Impact of Patient Demographics and Geographic Location. Acad Pathol. 2018;5. 10.1177/2374289518797573

18. Johnson C, Richwine C, Patel V. Individuals’ access and use of patient portals and smartphone health apps, 2020 [Internet]. HealthIT.gov; 2021 Sep. Available from: https://www.healthit.gov/data/data-briefs/individuals-access-and-use-patient-portals-and-smartphone-health-apps-2020

19. Hoogenbosch B, Postma J, de Man-van Ginkel JM, Tiemessen NA, van Delden JJ, van Os-Medendorp H. Use and the users of a patient portal: Cross-sectional study. J Med Internet Res. 2018;20(9). 10.2196/jmir.9418

20. Dumitrascu AG, Burton MC, Dawson NL, Thomas CS, Nordan LM, Greig HE, et al. Patient portal use and hospital outcomes. J Am Med Inform Assoc. 2017;25(4):447–53. 10.1093/jamia/ocx149

21. Sun R, Burke LE, Saul MI, Korytkowski MT, Li D, Sereika SM. Use of a patient portal for engaging patients with type 2 diabetes: Patterns and prediction. Diabetes Technol Ther. 2019;21(10):546–56. 10.1089/dia.2019.0074

22. Woods SS, Forsberg CW, Schwartz EC, Nazi KM, Hibbard JH, Houston TK, et al. The association of patient factors, digital access, and online behavior on sustained patient portal use: A prospective cohort of enrolled users. J Med Internet Res. 2017;19(10). 10.2196/jmir.7895

23. Rodriguez-Hart C, Gray I, Kampert K, White M, Wolfe C, Wilson M, et al. Just text me! Texting sexually transmitted disease clients their test results in Florida, February 2012-January 2013. Sex Transm Dis. 2015;42(3):162–7. 10.1097/OLQ.0000000000000242

24. Nyatsanza F, McSorley J, Murphy S, Brook G. ‘It’s all in the message’: The utility of personalised short message service (SMS) texts to remind patients at higher risk of STIs and HIV to reattend for testing—a repeat before and after study. Sex Transm Infect. 2015;92(5):393–5. 10.1136/sextrans-2015-052216

25. Lalla-Edward ST, Mashabane N, Stewart-Isherwood L, Scott L, Fyvie K, Duncan D, et al. Implementation of an mHealth App to Promote Engagement During HIV Care and Viral Load Suppression in Johannesburg, South Africa (iThemba Life): Pilot Technical Feasibility and Acceptability Study. JMIR Form Res. 2022;6(2):e26033. 10.2196/26033

